# A long-term prospective cohort study of seriously injured older trauma patients

**DOI:** 10.1101/2022.08.09.22278578

**Authors:** Sarah Ibitoye, Lily Bridgeman-Rutledge, Ben Carter, Philip Braude

## Abstract

A protocol for a prospective observational cohort study to investigate the effect of frailty on long-term outcomes in older adults admitted with trauma. Patients aged 65 years and older admitted to the Severn Major Trauma Centre at North Bristol NHS Trust in England between November 2018 and September 2019, will be followed up at 4-years. The objective is to determine if there is an association between Clinical Frailty Scale and mortality at 4 years after admission to hospital with a traumatic injury. The primary outcome will be mortality as measured by time from hospital admission to death. Analyses will be adjusted for other factors shown to be associated with mortality, using a mixed-effects multivariable Cox proportional hazards model.

**Protocol Version:** 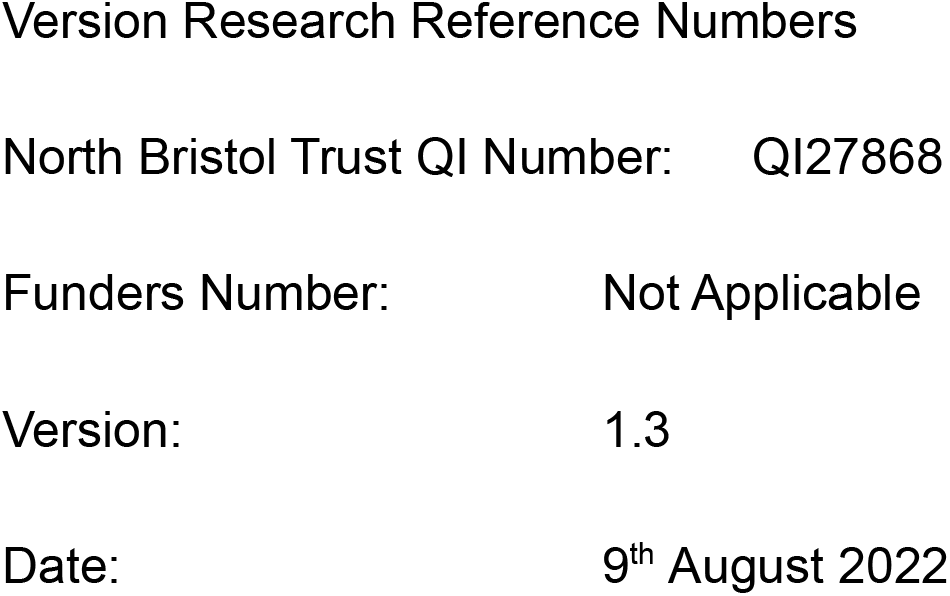

**Signature Page:** The undersigned confirm that the following protocol has been agreed and accepted and that the Chief Investigator agrees to conduct the study in compliance with the approved protocol and will adhere to the principles outlined in the Declaration of Helsinki, the Sponsor’s SOPs, and other regulatory requirements.

I agree to ensure that the confidential information contained in this document will not be used for any other purpose other than the evaluation or conduct of the investigation without the prior written consent of the Sponsor

I also confirm that I will make the findings of the study publicly available through publication or other dissemination tools without any unnecessary delay and that an honest, accurate and transparent account of the study will be given; and that any discrepancies from the study as planned in this protocol will be explained.

**Chief Investigator:** 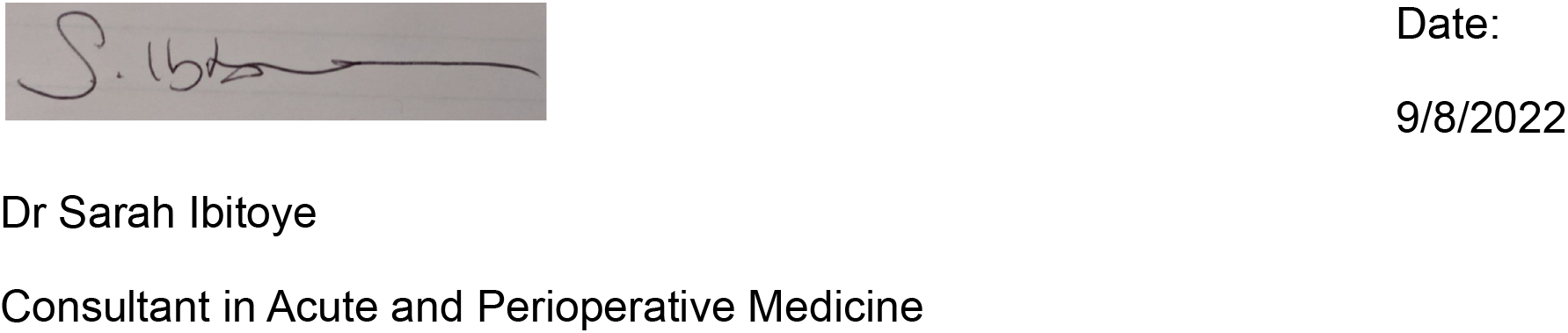

**Key Study Contacts:** 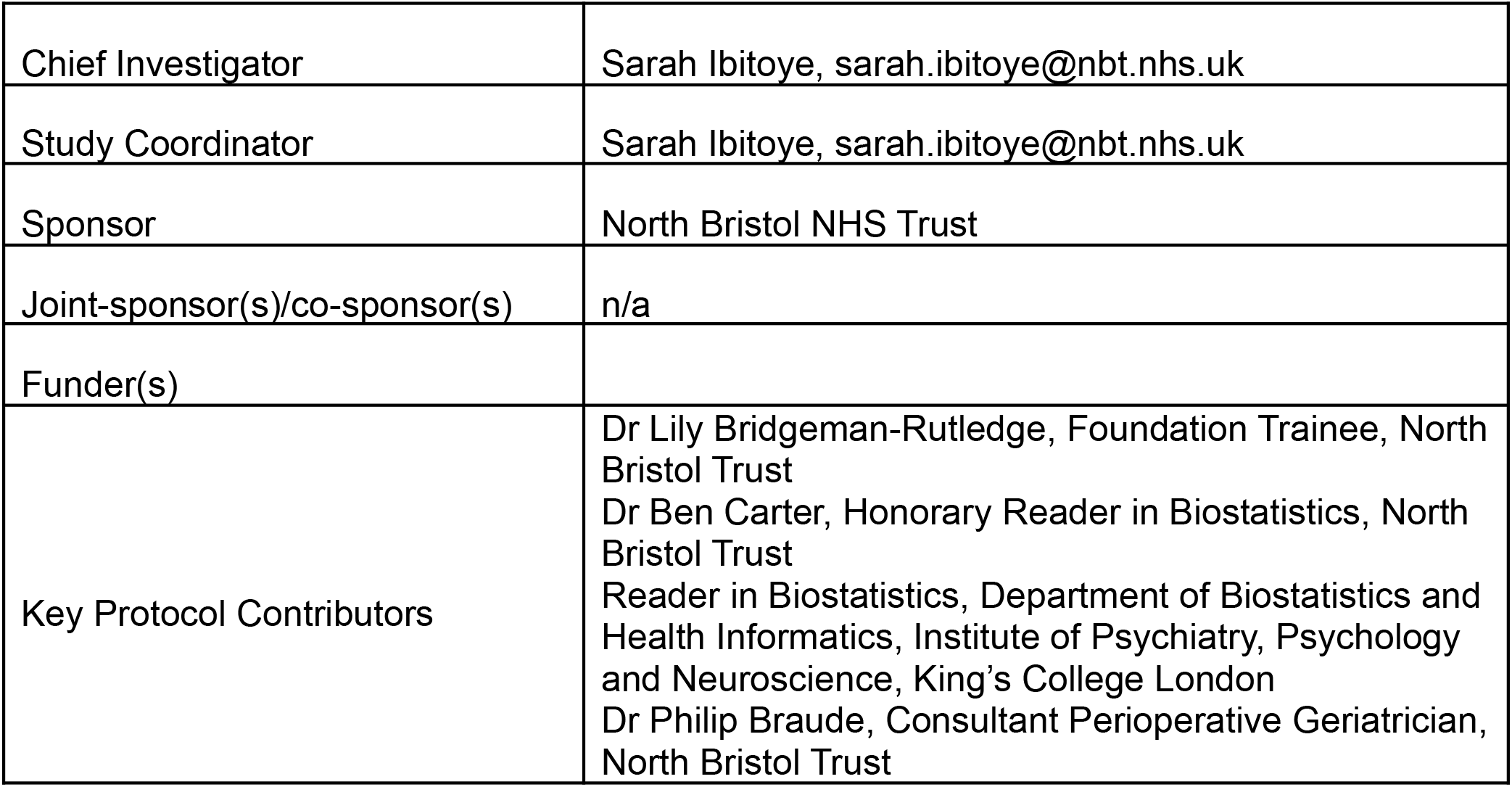

**Study Summary:** 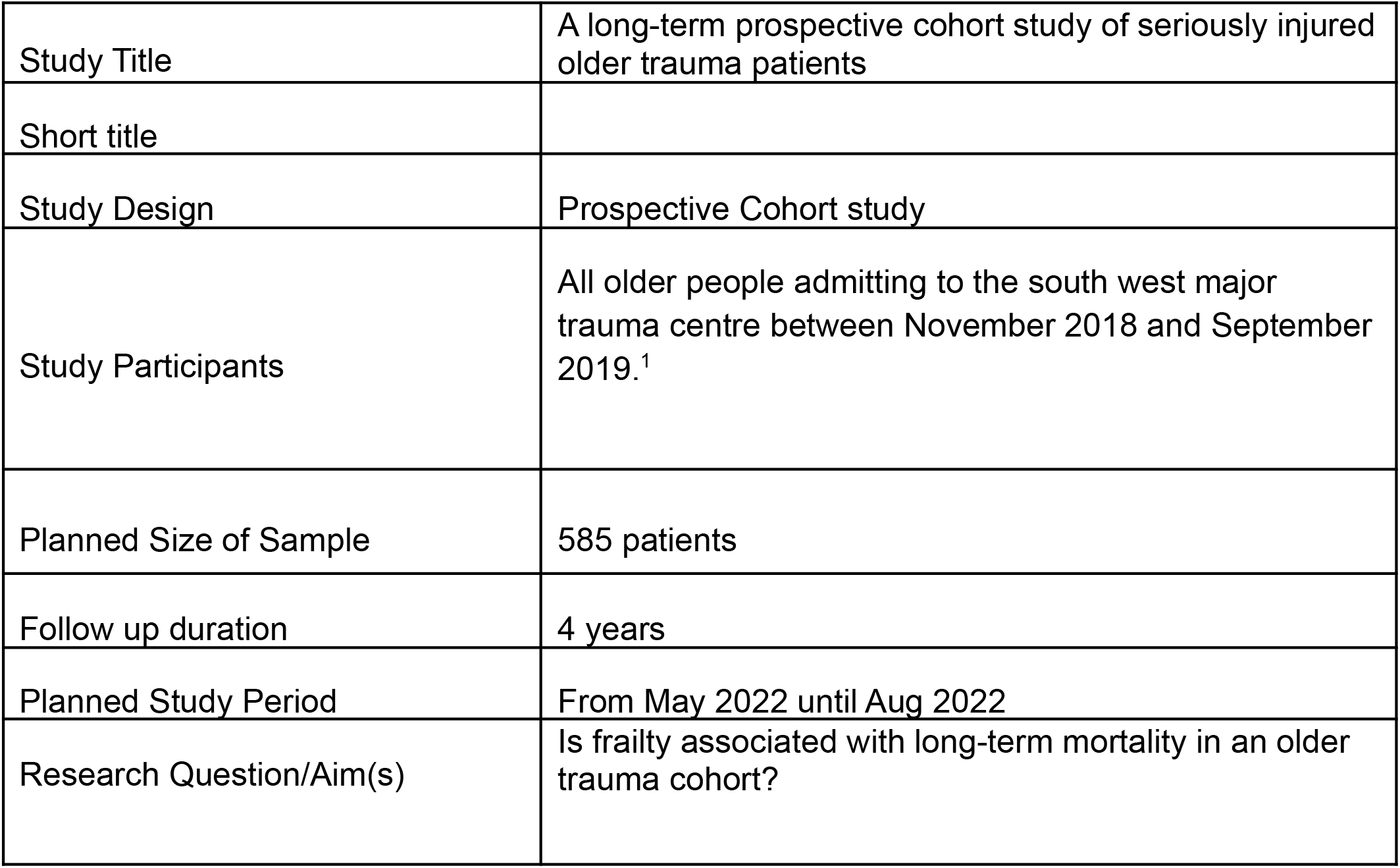

**Funding and Support in Kind:** 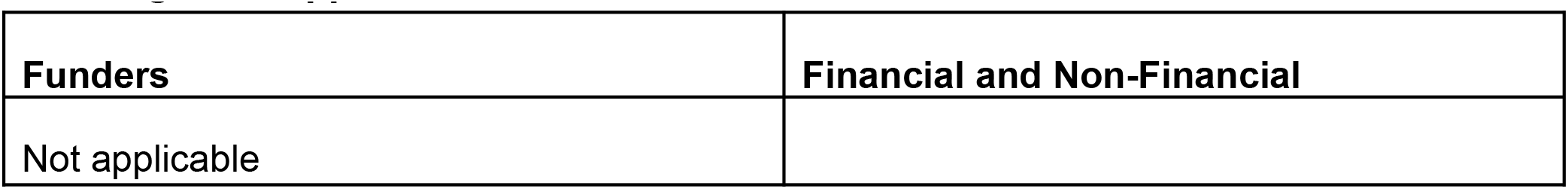

**Roles and Responsibilities:** *Protocol Contributors:* 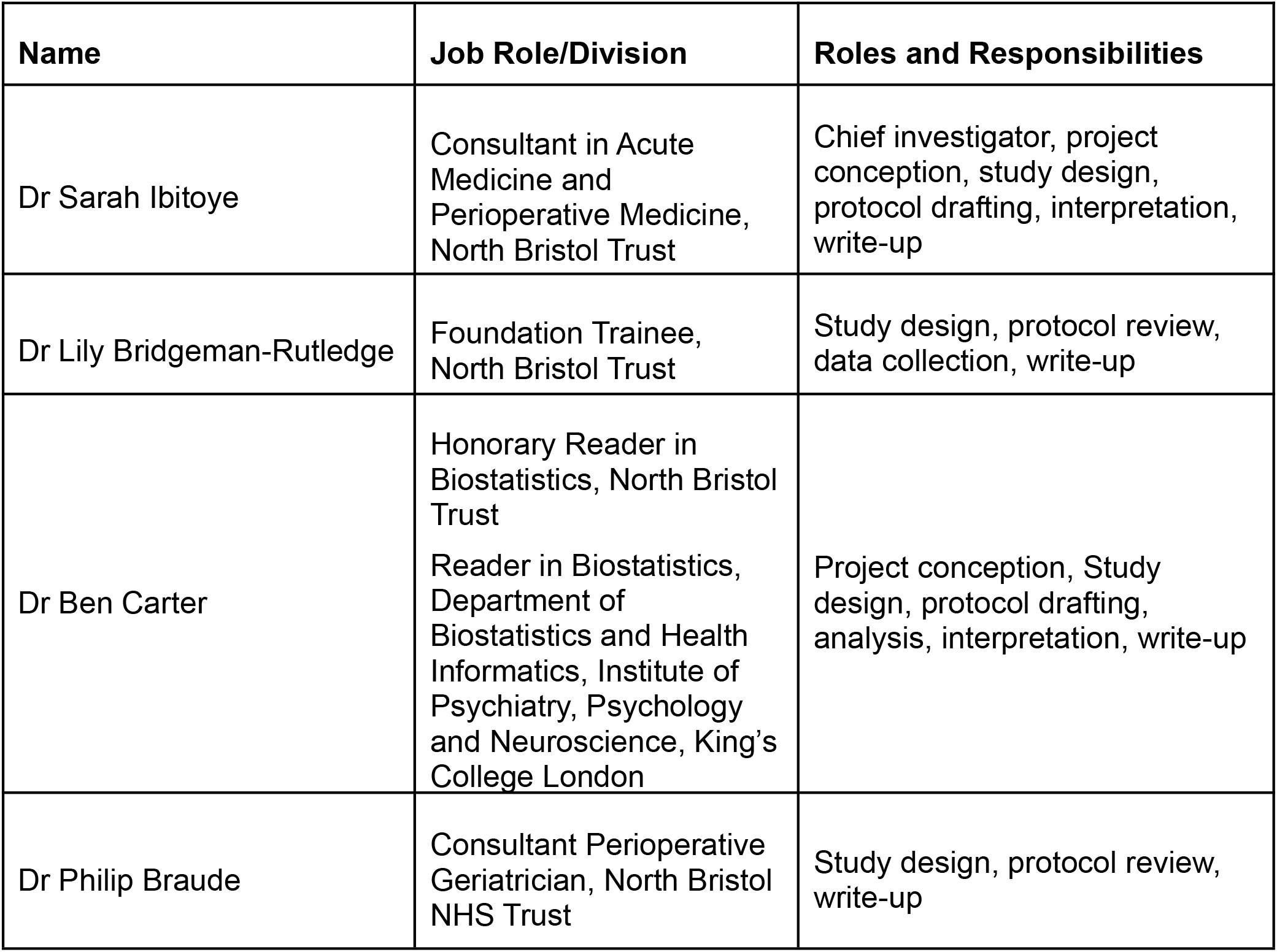

**Study Flow Chart:** 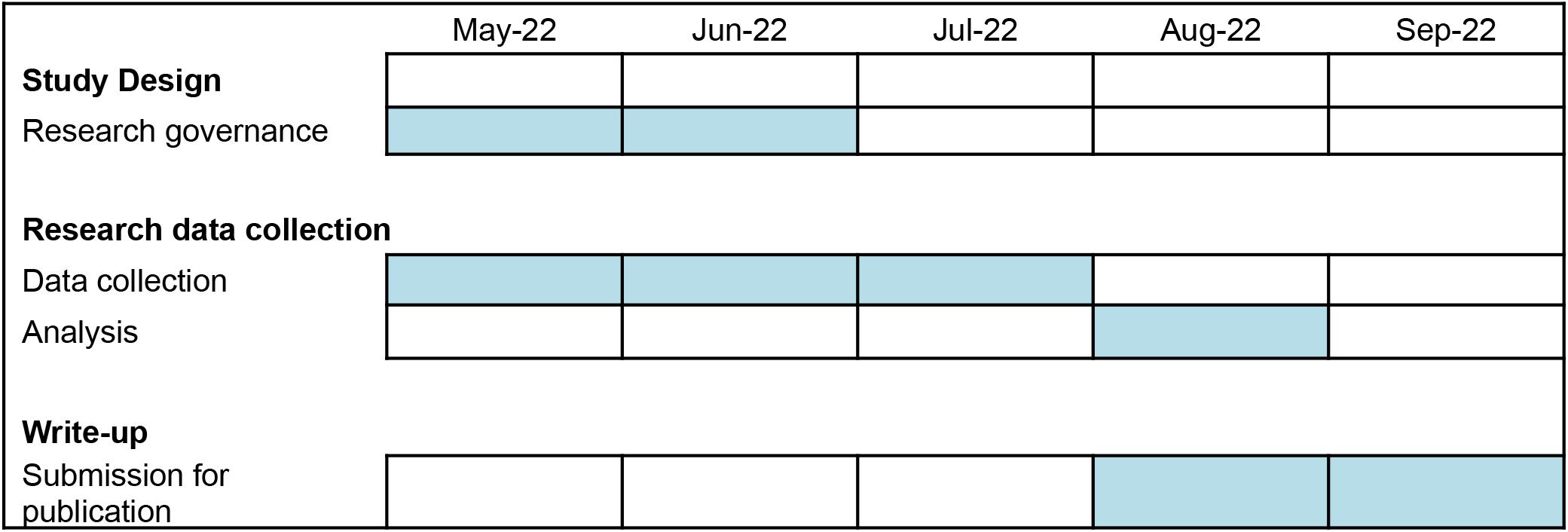

## 1 Background

Around half of United Kingdom (UK) major trauma admissions are adults aged 65 years or older.^2,3^ In this population many patients are living with frailty.^4^ Since 2019, frailty assessment has been a mandated part of clinical assessment in older adults admitted with major trauma, when it became part of the best practice tariff (BPT).^5^ Since then pre-admission frailty has been found to offer greater utility than age to predict short term mortality^4^, mortality up to 1-year;^1^ delirium^6^ and discharge destination.^4,7^

However, there remains a paucity of published data describing long-term outcomes for trauma populations, or defining predictors of mortality beyond 1-year. Pizzonia et al. found an association between frailty and long-term mortality (median follow-up 2.4 years) in hip fracture patients,^8^ however, it is unclear if this can be extrapolated to all trauma patients. In other populations frailty has been associated with all-cause mortality at 3-,^9^ 5-^10^ and 10-years.^11^ However, these findings were based on either community data,^10,11^ or patients admitted with acute medical conditions.^9^ In addition, these retrospective studies utilised electronic data to derive frailty indices, which may be less sensitive and specific than a prospective clinical frailty assessment in predicting long-term mortality.

As our older trauma population grows, tools to help predict long-term outcomes will be key in service planning and provision. Locally, an understanding of mortality for these individuals can help frame communication with patients and general practitioners at the time of hospital discharge. This may help to ensure that processes such as the Gold Standards Framework are utilised to improve care for people coming towards the end of their life.

Within North Bristol NHS Trust, data on older adults admitted with trauma was collected from November 2018 to September 2019. This was published showing an association between Clinical Frailty Scale (CFS) and mortality at 1-year.^1^ We can utilise this existing data to look at the effect of CFS on long-term mortality (up to 4-years) in our local trauma population.

## 2 Research Question

Is frailty associated with long-term mortality in an older trauma cohort?

### 2.1 Objectives

To determine if there is an association between the CFS and mortality at 4 years, in adults aged 65 and over admitted to hospital with a traumatic injury.

### 2.2 Outcomes

Mortality (time from hospital admission to death).

## 3 Study Setting

North Bristol NHS Trust hosts the Major Trauma Centre (MTC) for the Severn Major Trauma Network serving a population of 2.5 million adults in South West England. The MTC receives patients both directly and as transfers from eight surrounding trauma units. At the MTC a Geriatric Perioperative Care (G-POC) team was established in April 2019, to proactively deliver comprehensive geriatric assessment to older adults admitted with trauma. Prior to this patients were only seen by a geriatrician if a referral was made.

## 4 Study Design and Methods of Data Collection

### 4.1 Study design

A Prospective Cohort study.

### 4.2 Data Collection

Baseline data will be those collected for a previous study “Predicting 1 year mortality after traumatic injury using the clinical frailty scale”.^1^

Data were collected by the G-POC team retrospectively from November 2018 to March 2019, and prospectively from March to September 2019. A 1-month interruption to data collection occurred when the new G-POC team was being established.

The TARN database was used to extract patient demographics, injury pattern, and injury severity score (ISS). CFS was either retrospectively scored by a geriatrician from the hand-written notes which had been scanned on to the electronic patient record,^12^ or prospectively scored and entered directly into the database. All members of the G-POC team completed online CFS training to standardise scores.^13^

Mortality at 4 years post admission will be identified by review of NHS primary and secondary care linked electronic health records. For time-to-mortality analysis the last date known alive will be recorded as the date of any interaction between the patient and the NHS including an appointment, admission, or medication issue. Patients discharged out of the local area will be censored at the point of hospital discharge. Last date of follow-up was 8th July 2022.

#### Primary outcome

This will be the time from index hospital admission to mortality. Patients lost to follow up will be censored at the date last known alive.

#### Key exposure

This will be the clinical frailty scale (CFS) and analysed as CFS 4, 5, 6, 7-8, versus CFS 1-3.

### 4.3 Variables

The primary outcome will be time-to-mortality from hospital admission. Patient demographics and injury details will be collected including: age, gender, comorbidities using and Ageless Charlson Comorbidity Index, frailty using the CFS, injury type and injury severity score (ISS).

### 4.4 Data Storage

Data will be recorded directly into a bespoke spreadsheet. Electronic data will be stored securely at the local site. Access to the data will be by the study team only through a password protected file. Data will be processed at King’s College London

Data will be kept for 5 years after the study closes in case of external requests to verify published material. A unique key will be used to identify patients via a study ID number only.

### 4.5 Data Analysis

Demographics and baseline clinical characteristics will be presented against long term survival. A Kaplan Meier plot will be presented to visually assess the baseline proportionality and survival curves for those that are frail, versus those that are not frail, with included at risk table.

The primary outcome of mortality will be assessed as the time-to-mortality with a multivariable Cox proportional baseline hazards model to assess the effect of frailty (CFS 1-8) on mortality.

The analysis will be adjusted for factors previously shown to be associated with mortality in this population,^1,3^ including: age group, gender, time-period, ISS, major post-injury complications, most severely injured body part, if underwent surgery (Y/N). Both crude hazard ratio (HR) and adjusted hazard ratio (aHR) will be estimated with associated 95% confidence intervals (CI). Baseline proportionality will be assessed visually using log-log residual plots.

Data will be analysed in Stata version 17.0.

#### Subgroup Analysis

We plan to estimate the difference between those that are frail and not frail (CFS 1-3, versus CFS 4-8) in the following subgroup: Age group; sex; time-period; ISS; surgery; and complications

Missing data will be explored using pattern missingness and presented within the demographics and index hospital admission.

## 5 Sample and Recruitment

In previous work we identified a non-significant hazard ratio of 1.8 between CFS 4, versus CFS 1-3^1^. In order to detect this difference with 80% power and a 5% significance level we would need 45 events within each group. Previously we identified 147 events from 585 patients admitted at 1-year and we estimate that there will be 200 events after 4 year follow up.

## 6 Ethical and Regulatory Considerations

### 6.1 Assessment and management of risk

Data security issues: data will be stored on Trust computer systems with password protection. Only those in the study team will have access to these data.

Identifiable patient information issues: analysis will take place using anonymised records. Patient level and hospital level identifiers will be removed.

### 6.2 Ethical approval

The study has been registered with the Quality Assurance and Clinical Audit Department at North Bristol NHS Trust (ID27868). Ethical review was not deemed to be required as all data were collected as part of routine clinical practice.

### 6.3 Peer review

This protocol has been reviewed by the chief investigator and study contributors.

### 6.4 Access to the Final Study Dataset

In line with many peer reviewed journal’s policy for data sharing, data sharing may be offered to third parties only on request to the study Chief Investigator. Data will be shared with anonymous records if deemed appropriate, arranged via data sharing agreement, and transferred using secure systems.

## Data Availability

All data produced in the proposed study may be available upon reasonable request to the chief investigator.

## 7 Dissemination Policy

### 7.1 Dissemination policy

The data will be owned by North Bristol Trust as the sponsor. On completion of the study the data will be analysed and a study report completed.

It will be disseminated locally via the trust operational update and events, as well as more widely through national and international conference presentations. A summary of the work will be submitted for peer-reviewed publications.

### 7.2 Authorship Eligibility Guidelines

Authorship will take into account all persons involved in the study design, analysis and write up. This will be accurately reflected when any papers are submitted for peer-reviewed publication.

## Notes

### Competing Interest Statement

The authors have declared no competing interest.

### Funding Statement

This study did not receive any funding.

### Author Declarations

Quality Assurance and Clinical Audit Department of North Bristol NHS Trust waived ethical approval for this work.

